# Is mRankin scale correlated with mTICI? A systematic review and meta-regression on RCTs and registries

**DOI:** 10.1101/2021.05.12.21257077

**Authors:** Gianluca De Rubeis, Enrico Pampana, Luca Prosperini, Sebastiano Fabiano, Luca Bertaccini, Sabrina Anticoli, Luca Saba, Claudio Gasperini, Enrico Cotroneo

**Affiliations:** Department of Diagnostic, UOC of Diagnostic and Interventional Neuroradiology, San Camillo-Forlanini Hospital, Rome, Italy; Department of Neuroscience, UOC Neurology, S Camillo Forlanini Hospital, Rome Italy; Emergency Department, UOSD Stroke Unit, S. Camillo-Forlanini Hospital, Rome, Italy; Department of Medical Imaging, Azienda Ospedaliero Universitaria (A.O.U.) of Cagliari-Polo di Monserrato, Cagliari, Italy

**Author notes:** Co-first author, equally contribution to the manuscript (Gianluca De Rubeis, Enrico Pampana).

**Keywords:** Stroke, Cerebral Infarction, Randomized Controlled Trial, Observational Study, Mechanical Thrombolysis

## Abstract

**Background and Purpose:** mTICI ≥2b/3 is one of the strongest positive predictors of mRS ≤2. Quantitative analysis is poorly investigated. Reconcile results from RCT and registries is still a challenge.

The purpose was to evaluate the numeric correlation between mTICI≥2b/3 and mRS≤2 in RCT and registries.

**Methods:** Literature research was performed on Pubmed for studies in 2015-2020. mTICI, mRS and sample size were recorded. Exclusion criteria were monocentric study, not-human and not-English. Studies quality were assessed with MINORS and RoB2. Meta-logistic and meta-linear regressions were used to correlate mTICI and mRS in both RCTs and registries. Z-test was used for comparing coefficients between RCTs and registries.

**Results:** Twenty-six studies were evaluated (13 registries; 14 RCTs) for 24423 patients (21914 from registries [average per registry 1685±1277]; 2509 from RCTs [average per RCT 179±160]). RCTs involved anterior circulation only, 7/13 (53.8%) registries considered also posterior one.

The OR of obtaining a mRS≤2 for a singular increased of mTICI ≥2b rate was 1.65 (CI95% 1.22-2.01) for all studies, 1.65 (CI95% 1.10-2.46) for RCTs and 1.50 (CI95% 1.00-2.23) for registries. mTICI≥2b and mRS had a positive correlation with a coefficient of 0.49 (CI95% 0.19-0.80, p=0.001) for all studies, 0.54 (CI95% 0.09-1.00) for RCTs and 0.42 (CI 95% 0.04-0.81) for registries. No differences were found in the coefficients between RCTs and registries (p=0.63; p=0.65; respectively).

**Conclusions:** Unitary increased of mTICI≥2b rate correspond to an augment of mRS≤2 by 0.50 (CI95% 0.19-0.89) with OR of obtaining mRS≤2 of 1.65 (CI95% 1.22-2.01), without significantly differences in coefficients.

## Manuscript

## Introduction

Since the publication in 2015 of the successful 5 randomized controlled trials (RCT)[1] on the endovascular mechanical thrombectomy, the new era of stroke treatment has begun. In the following years, a submergent literature has been published including RCTs and registries. mRankin scale (mRS) at 90 days is the functional primary endpoint in all RCTs and, generally, it is dichotomized into 0-2 (good functional outcome) and ≥3[1 2]. However, from procedural side, the goal is modified thrombolysis in cerebral infarction (mTICI) of ≥2b[3] (successful revascularization). Although, a positive correlation between mTICI v and mRS≤2 is well established[4-6], a numerical analysis is poorly investigated.

RCTs is the gold standard for medical research, however registries could represented the “real-world” clinical practise[7]. A reconciliation between results originating from RCTs and registries is still a challenging in medicine[8]. Although, a Cochrane’s systematic review[7] showed little differences in healthcare outcomes between these two types of study (rate of odds ratio: 1.08), Deb-Chatterji et al[9] demonstrated a worst performance for functional independency and mortality in stroke registry data comparing with the “first five” trials and HERMES meta-analysis data.

The aim of the present study was to evaluate the numerical correlation between successful recanalization (mTICI≥2b/3) and good functional outcome (mRS≤2) in both RCTs and registries using meta-regressions.

## Methods

The study protocol was available upon request by mailing the corresponding author. The data used for the systematic review and the meta-regressions were publicly available.

Literature research in Pubmed was performed for published multicentric RCTs and registries, from 2015 (first publication of successful RCT in endovascular stroke) to 2020, on mechanical thrombectomy in stroke. The keywords were “stroke”, “thrombectomy”, “randomize controlled trial” and “registry”. The inclusion criteria for the studies were: English literature, humans, and mechanical thrombectomy in at least one of the study’s arm. Title and abstract were reviewed for considering inclusion. Further the full text was analysed. Monocentric studies were excluded. For ASTER[10] and COMPASS[11] trials, which compared contact aspiration and stent retriever, the data were considered unitary. For DIRECT-MT trial[12] which confronted endovascular treatment with or without Alteplase administration, the data were considering unified. The detailed flowchart was described in **Figure 1**.

**Figure 1.**
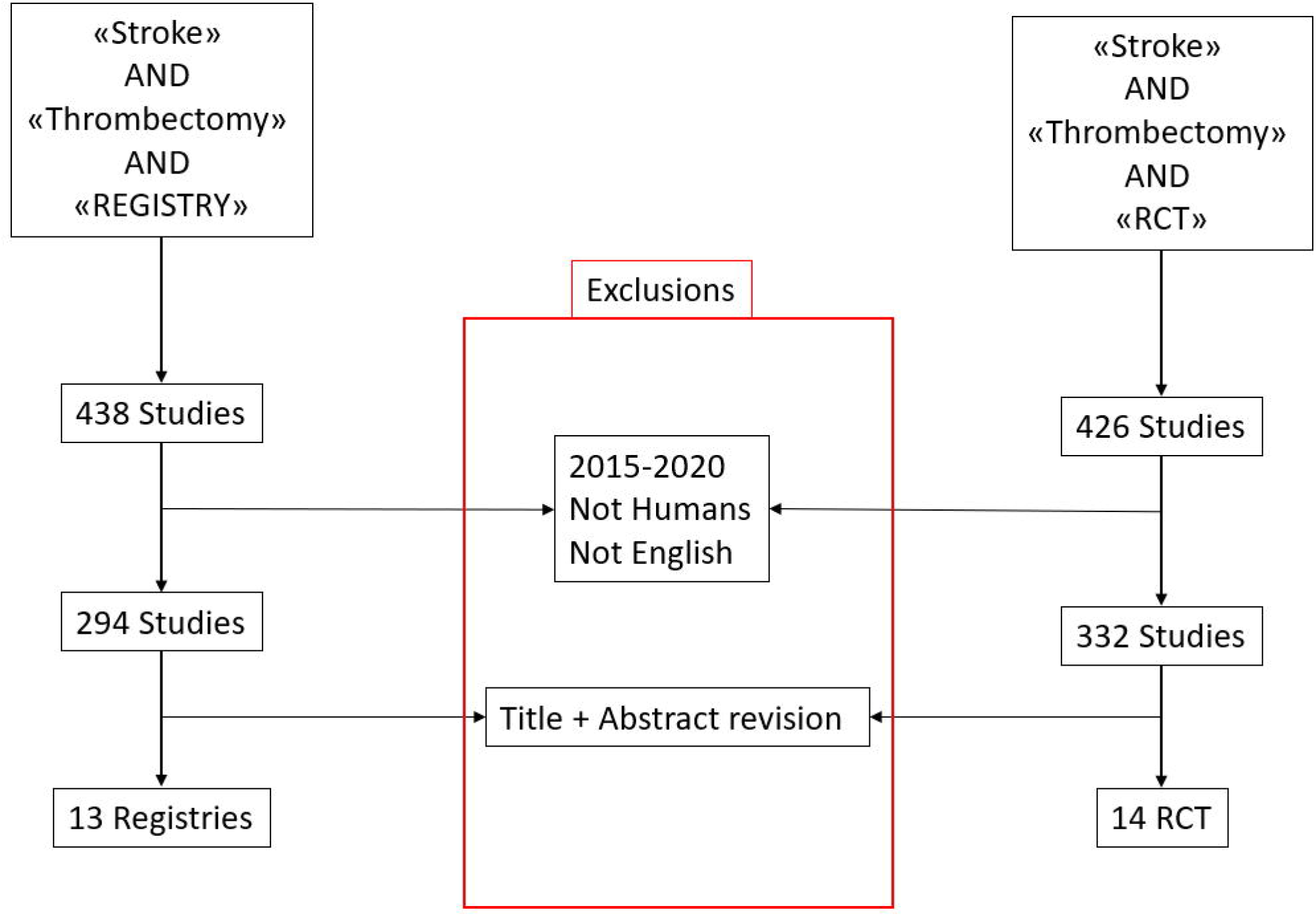
Study flowchart

### Study quality assessment

Studies quality was assessed with methodological index for non-randomized studies (MINORS) criteria[13] for registry and with Risk of Bias 2 (RoB2) tool[14] for RCT. The results of the assessment were described in **Table 1**.

**Table 1.** List of the studies.

| **Name** | **Type** | **Trial type** | **Year** | **Risk of Bias** | **Number of patients** | **Comparison for RCT**<br>**Non-comparison for Registry** | **Inclusion criteria** | **mTICI $\geq 2b$ (%)** | **90 days mRS $\leq 2$ (%)** |
| --- | --- | --- | --- | --- | --- | --- | --- | --- | --- |
| **EXTEND-IA<sup>a</sup>** | RCT | Prospective, blind end-point evaluation | 2015 | Low | 35 | SR + Alteplase vs Alteplase alone | Alteplase within 4.5 hours from the onset; Anterior circulation | 86 | 71 |
| **REVASCAT** | RCT | Prospective, sequential, blind end-point evaluation | 2015 | Low | 103 | SR vs medical therapy | Treated within 8 hours from the onset; Anterior circulation | 65.7 | 44 |
| **MR CLEAN** | RCT | Prospective, pragmatic, blind end-point evaluation | 2015 | Low | 233 | SR or Thrombolytic Agent vs medical therapy | Treated within 6 hours from the onset; Anterior circulation | 58.7 | 33 |
| **ESCAPE** | RCT | Prospective, blind end-point evaluation | 2015 | Low | 165 | SR vs medical therapy | Treated within 12 hours from the onset; Anterior circulation | 72.4 | 53 |
| **SWIFT-PRIME** | RCT | Prospective, blind | 2015 | Low | 98 | SR + Alteplase vs Alteplase alone | Treated within 6 | 88 | 60 |

|  |  | end-point evaluation |  |  |  |  | hours from the onset; Anterior circulation |  |  |
| --- | --- | --- | --- | --- | --- | --- | --- | --- | --- |
| **Therapy<sup>a</sup>** | RCT | Prospective | 2016 | Low | 55 | CA vs medical therapy | Eligible for alteplase; Anterior circulation; NIHSS $\geq 8$ | 70 | 38 |
| **THRACE** | RCT | Prospective, blind end-point evaluation | 2016 | Low | 204 | Mechanical thrombectomy + alteplase vs alteplase | Treated within 5 hours from the onset; Anterior circulation | 69 | 53 |
| **ASTER** | RCT | Prospective, blind end-point evaluation | 2017 | Low | 381 | CA vs SR | Treated within 6 hours from the onset; Anterior circulation | 84.8 | 47.6 |
| **PISTE<sup>a</sup>** | RCT | Prospective, blind end-point evaluation | 2017 | Low | 33 | Mechanical thrombectomy + alteplase vs alteplase | Eligible for alteplase; Anterior circulation; | 87.4 | 51 |
| **STRATIS** | Registry | Prospective | 2017 | 13 | 989 | SR (Solitaire) | Treated within 8 hours from the onset;<br>Anterior and<br>Posterior<br>circulation | 87.9 | 56.5 |
| **SITS-TBY** | Registry | Prospective | 2017 | 12 | 1053 | Mechanical thrombectomy | European guidelines;<br>Anterior circulation | 74 | 48 |
| **ACTUAL** | Registry | Retrospective | 2018 | 11 | 698 | Mechanical thrombectomy | AHA/ASA indications;<br>Anterior circulation | 83.8 | 43.8 |
| **Trevo 2000** | Registry | Prospective | 2018 | 14 | 1192 | SR (TREVO) | Center-based indications | 92.8 | 55.3 |
| **MR CLEAN** | Registry | Prospective | 2018 | 11 | 1628 | Mechanical thrombectomy | Treated within 6.5 hours from the onset;<br>Anterior circulation | 58.7 | 37.9 |
| **DEFUSE 3** | RCT | Prospective, blind end-point evaluation | 2018 | Low | 92 | Mechanical thrombectomy | 6-16 hours from the onset;<br>imaging mismatch;<br>anterior circulation | 76 | 45 |
| **ETIS** | Registry | Prospective | 2019 | 13 | 1541 | Mechanical thrombectomy | AHA/ASA indications; Anterior and posterior circulations | 76.6 | 44.1 |
| **COMPASS** | RCT | Prospective, blind end-point evaluation | 2019 | Low | 270 | CA vs SR | Treated within 6.5 hours from the onset; Anterior circulation | 80.7 | 50.4 |
| **Track** | Registry | Retrospective and prospective | 2019 | 13 | 624 | SR (TREVO) | AHA/ASA indications; Anterior and posterior circulations | 80.7 | 48.3 |
| **BEYOND-SWIFT** | Registry | Retrospective | 2019 | 11 | 2046 | Mechanical thrombectomy | AHA/ASA indications; Anterior and posterior circulations | 71.4 | 45.5 |
| **German** | Registry | Prospective | 2019 | 13 | 2794 | Mechanical thrombectomy | AHA/ASA indications; Anterior and posterior circulations | 83 | 37 |
| **Moscow** | Registry | Retrospective | 2019 | 12 | 742 | Mechanical thrombectomy | AHA/ASA indications; Anterior and posterior circulations | 75 | 29.9 |
| **STAR** | Registry | Unknown | 2020 | 12 | 3850 | Mechanical thrombectomy | AHA/ASA indications; Anterior and posterior circulations | 84 | 41 |
| **IRETAS** | Registry | Retrospective | 2020 | 13 | 4429 | Mechanical thrombectomy | AHA/ASA indications; Anterior and posterior circulations | 75.1 | 43.7 |
| **Resiliet<sup>a</sup>** | RCT | Prospective | 2020 | Low | 111 | Mechanical thrombectomy vs standard care | AHA/ASA indications; Anterior circulation | 82 | 35.1 |
| **TREAT** | Registry | Prospective | 2020 | 11 | 328 | Mechanical thrombectomy | AHA/ASA indications; Anterior and posterior circulations | 83.3 | 43.9 |
| **DIRECT MT** | RCT | Prospective | 2020 | Low | 622 | Mechanical thrombectomy vs Mechanical thrombectomy+Alteplase | AHA/ASA indications; Anterior r<br>circulati<br>on | 82.0 | 47.3 |

SR: stent retriever; CA: contact aspiration; × Contact aspiration; °Stent retriever; α interrupted early; NIHSS: National Institute Health Stroke Scale

### Data extraction, statistical analysis, and results interpretation

For each study, the percentage of successful revascularization (mTICI≥2b), rate of good functional outcome at 90 days (mRS≤2) and the number of patients enrolled were recorded (Table 1.). Data were divided into two categories: RCT and registry. The data extraction were performed by two interventional neuroradiologist in consensus (BLIND and BLIND, 4 and 5 years of experiences, respectively).

The data were analysed using meta logistic regression analysis with random effect model, using as covariate: mTICI≥2b, studies involved also posterior circulation and types of studies (RCTs and registries). Since, the mRS percentage across the studies are continuous data and follow a normal distribution (Kolmogorov-Smirnov test), these data respect also the assumption of central limit theorem to perform a meta linear regression analysis[15]. Both linear and logistic meta-regressions were performed. Z’s test proposed by Clogg et al[16] was used for comparing the coefficient of the two meta-regressions. p≤0.05 was considered as significant. R-Studio (R-project http://www.R-project.org) and OpenMeta version 12.11.14 (http://www.cebm.brown.edu/openmeta/index.html) were used as statistical software. The graphs were plotted with Microsoft Excel (Office 365, Microsoft Corporation, Redmond, USA).

Meta-regression was performed using both linear and logistic setting (according to the central limit theorem) due the different interpretation of the data resulted by the analysis. In particular, the coefficient of meta linear regression described the numerical correlation between mTICI percentage and 3-months mRS rate, more specifically the coefficient corresponds to how much the percentage of mRS≤2 increases per single increase in mTICI≥2b rate. On the contrary the odd ratio, derived from the meta logistic regression, represents the constant effect of a mTICI≥2b percentage, on the likelihood that mRS≤2 rate will occur; in the other words, the augmented chances of obtaining mRS≤2 per single increase of mTICI≥2b percentage.

### Study’s Outcomes

The primary outcome was to evaluate the correlation and the numeric relationship between successful revascularization (mTICI≥2b) and good functional outcome at 90 days (mRS≤2).

The secondary outcome was to observe if the correlation and the numeric relationship varied between RCTs and registries.

## Results

### Study population

The final population encompassed 27 studies (14 registries and 13 RCTs) for a total number of patients of 24423 divided into 21914 from registries (average per registry 1685±1277) and 2509 from RCTs (average per RCT 179±160) (**Flowchart 1**.). All RCTs were focused only anterior circulation while 7/13 (53.8%) of the registries considered also posterior one. (**Table 1**). A detailed list of inclusion criteria of the studies were described in **Table 1**.

### Results

The heterogeneity of the studies was high with an I^2^=92.8%, which decreased to I^2^=75.9% considering RCTs only and increased to I2=95.7% for registries.

The coefficient of the meta-logistic regression analysis for mTICI ≥2b was 0.50 (CI95% 0.20-0.70. p=0.001) for all studies, with a corresponding Odd-Ratio (OR) of 1.65 (CI95% 1.22-2.01) (**Figure 2**); no other covariates were significant (**Table 2)**. By performing meta logistic regression using mTICI and trial type as covariates, the coefficient for registries was -4.00 (CI95% -9.0-1.0, p=0.12) with an OR 0.018 (CI95% 0.0001-0.37). For RCTs, the coefficient was 0.50 (CI95% 0.10-0.90. p=0.02) with an OR of 1.65 (CI95% 1.10-2.46) and for registries was 0.40 (CI 95% 0.0-0.80 p=0.03) with an OR of 1.5 (CI95% 1.00-2.23). No differences were highlighted between the two coefficients (z=0.35; p=0.63) The coefficient of the meta-linear regression analysis for mTICI≥2b was 0.49 (CI95% 0.19-0.80, p=0.001) (**Figure 2**), by means that on average an increase of 1% of mTICI≥2b will increase the chance of obtaining a good functional outcome (mRS ≤2) by 0.49%; no other covariates were significant (**Table 3**.). After performing the meta linear regression using as covariates mTICI and trial type only, the coefficient for registries was -4.54 (CI95% -9.89-0.80, p=0.10). For RCTs, the coefficient was 0.54 (CI95% 0.09-1.00, p=0.02) and for registries was 0.42 (CI 95% 0.04-0.81, p=0.03). No differences were demonstrated between the two coefficients (z=0.39; p=0.65)

**Table 2.**
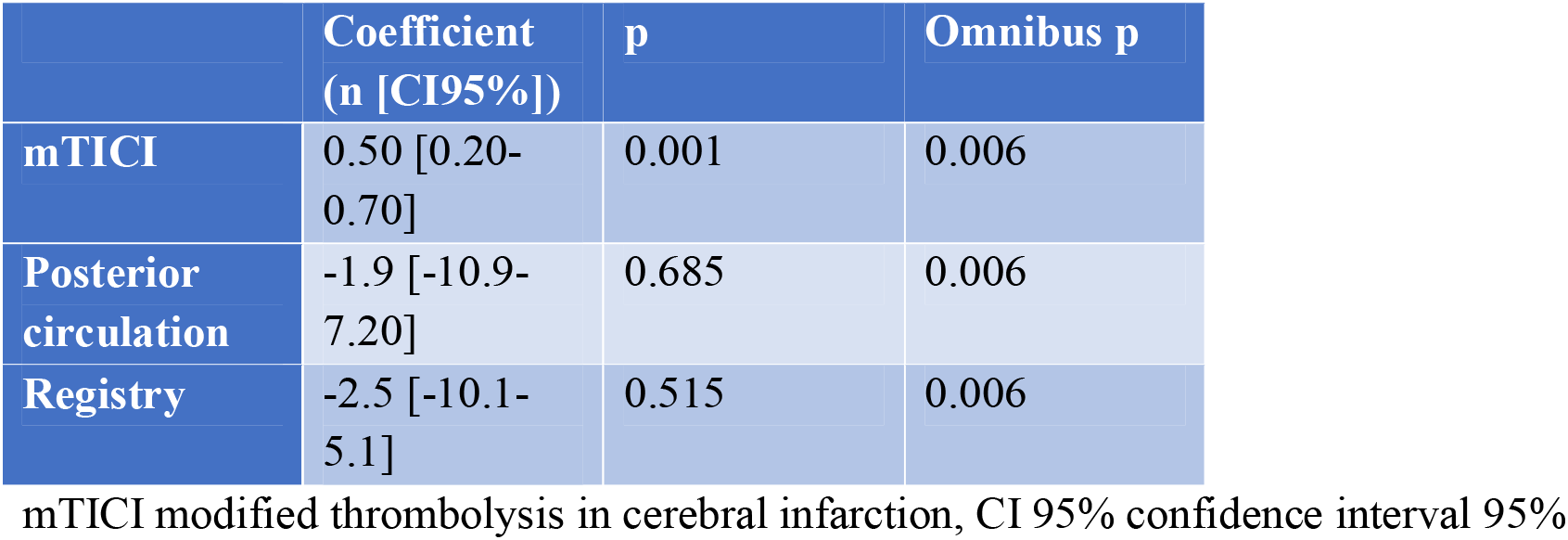
Meta logistic regression.

|  | Coefficient<br>(n [CI95%]) | p | Omnibus p |
| --- | --- | --- | --- |
| mTICI | 0.50 [0.20-0.70] | 0.001 | 0.006 |
| Posterior<br>circulation | -1.9 [-10.9-7.20] | 0.685 | 0.006 |
| Registry | -2.5 [-10.1-5.1] | 0.515 | 0.006 |

mTICI modified thrombolysis in cerebral infarction, CI 95% confidence interval 95%

**Table 3.**
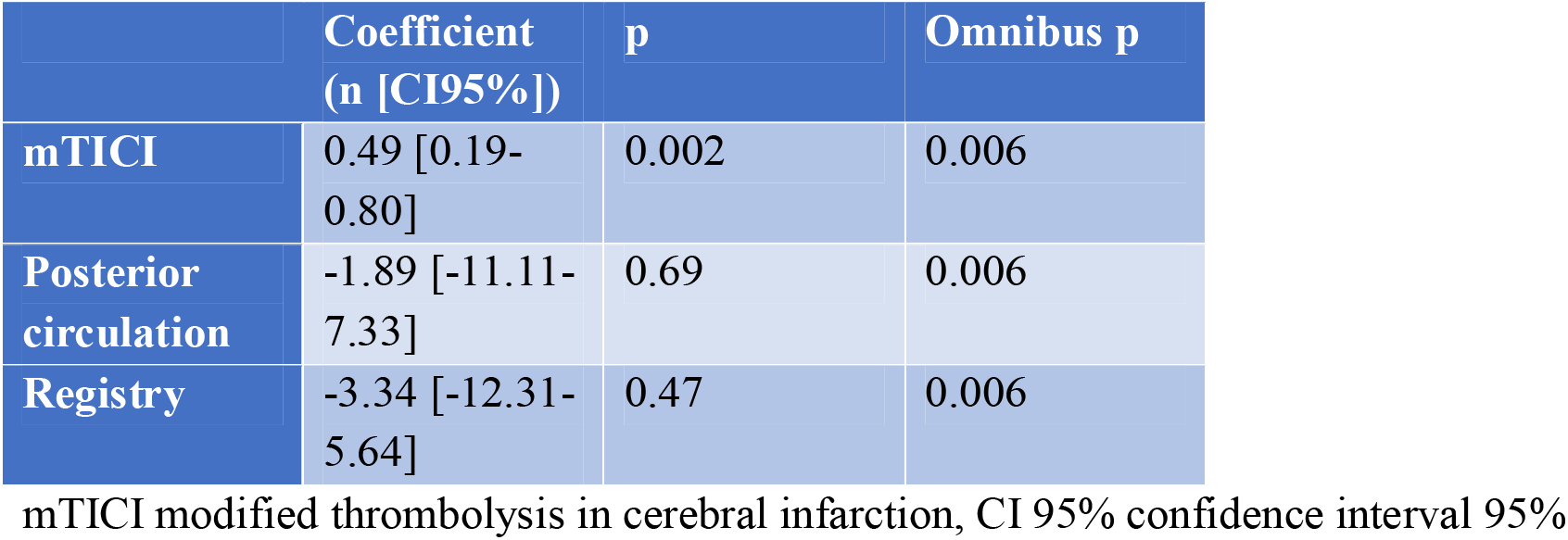
Meta linear regression.

|  | Coefficient<br>(n [CI95%]) | p | Omnibus p |
| --- | --- | --- | --- |
| **mTICI** | 0.49 [0.19-0.80] | 0.002 | 0.006 |
| **Posterior**<br>**circulation** | -1.89 [-11.11-7.33] | 0.69 | 0.006 |
| **Registry** | -3.34 [-12.31-5.64] | 0.47 | 0.006 |

mTICI modified thrombolysis in cerebral infarction, CI 95% confidence interval 95%

**Figure 2.**
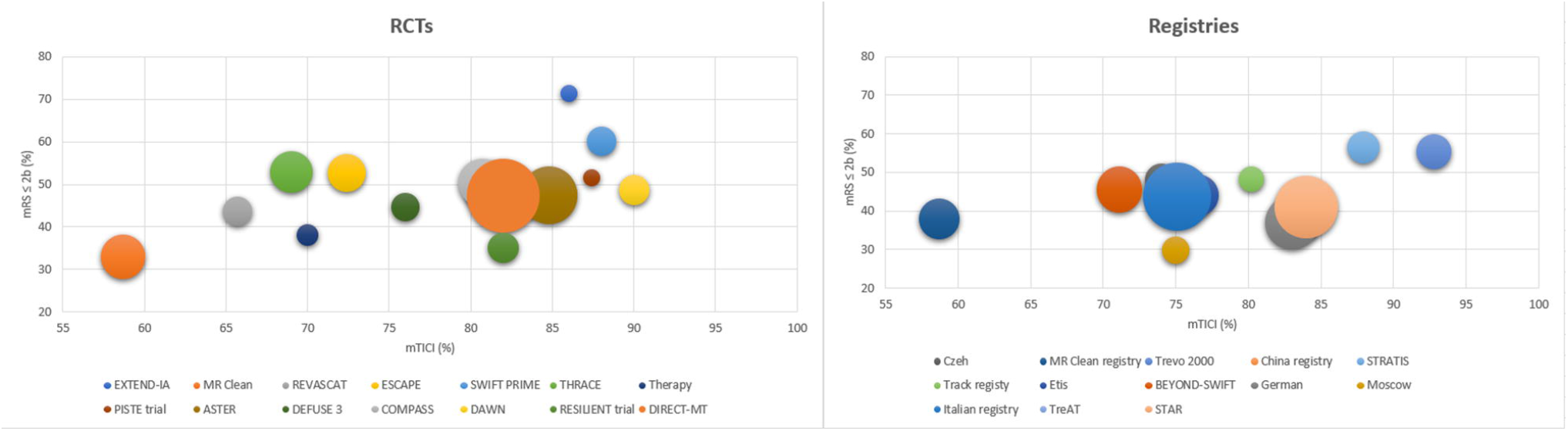
Bubble plot of the meta regression in RCTs (a) registries (b). As observed, the was a linear correlation between mTICI≥2b and mRS>2 using both analyses with a trending minor slope in registries. The size of the bubble represented the numerosity of the study

## Discussion

The OR of obtaining a functional independency (mRS≤2) for a unitary increased of mTICI ≥2b was 1.65 (CI95% 1.22-2.01) for all studies and 1.65 (CI95% 1.10-2.46) for RCTs and 1.5 (CI95% 1.00-2.23) for registries. A linear positive correlation was found between mTICI≥2b and mRS with a coefficient of 0.49 (CI95% 0.19-0.80, p=0.001) for all studies, 0.54 (CI95% 0.09-1.00) for the RCTs and 0.42 (CI 95% 0.04-0.81, p=0.03) for registries, without significant differences between the two coefficients (p>0.05).

Successful revascularization (mTICI≥2b) is considered one of the strongest predictor of functional independency (mRS≤2)[6], for this reason the AHA/ASA guidelines[3] addressed mTICI≥2b as the angiographic goal for maximizing the chance of obtaining good functional clinical outcome. A linear relationship between mTICI and mRS in RCTs was already observed; although, without numeric analysis[17 18]. The present meta-linear regression numerically confirmed the significantly correlation defining a comprehensive coefficient of 0.49 (CI95% 0.19-0.80, p=0.001). In addition, applying the meta-logistic regression the OR of obtain functional independency was 1.65 (CI95% 1.22-2.01).

Despite, RCTs is considered the gold standard for medical research, the problem of transporting results to “real-life world” is still an unsolved problem in medicine[19]. Basically, the internal validity of RCT may limit its external validity due to the different conditions and homogeneity of a RCT compared to the more complex and heterogeneous setting of a registry[20 21]. Moreover, performance bias is a matter of concern in both RCTs and registries, especially for non-pragmatic RCT in which the issues of “structured environment” may limited results generalization[22-24]. In addition, RCTs’ patients are generally strictly monitored and more sensitized on the disease comparing with observational registries[25]. In addition, mRS has several limitations including long time span between treatment and outcome assessment and lack of stroke-specificity disability[26]. In addition, mRS is influenced also by post-stroke care and rehabilitation[27-30]. The present data regarding mechanical thrombectomy in stroke partially confirmed these results. In fact, both the OR (meta-logistic regression) and the coefficient (meta-linear regression) were higher in RCT vs registries (OR: 1.65 [CI95% 1.10-2.46] vs 1.5 [CI95% 1.00-2.23], respectively) (coefficient: 0.54 [CI95% 0.09-1.00] vs 0.42 [CI 95% 0.04-0.81], respectively) (**Figure 2**.). These findings meaned that by increasing of 1% in mTICI≥2b rate the corresponding chance of augment of mRS ≤2 was 0.54 for RCTs and 0.42 registries. Namely, an angiographic success in mechanical thrombectomy has a higher impact on the clinical functional outcome if the patient is enrolled in a RCT despite in a registry. Despite its clinical importance, these differences remained not statistically significant in this present study due to small sample size. Interestingly, the lowest performance of mRS≤2 was reported by MR-CLEAN trial[31], the sole RCT who explicitly affirmed its pragmatic design. Under these lights, a direct comparison of a 3 months functional scale between real world (registries) and experimental setting (RCTs) is difficult to establish.

These discrepancies in clinical outcome may arise from several points. Firstly, RCTs and registries design (structured vs real world). Secondly, RCTs’ centres may be more expert in mechanical thrombectomy than registries’ one with a higher number of procedures per year which is a known prognostic factor[32]. Thirdly, the post-stroke care could be more effective in a high-volume centre. Fourthly, since a double-blind design is unfeasible, an assignment bias may impact on the results.

The current study presents several limitations. Firstly, the literature research was performed on PubMed and the authors did not have the access to original data. Secondly, the post-stroke care quality was not possible to assess, since was not reported in any papers. Thirdly, the studies presented different inclusion criteria which may slightly influence the enrolment. Fourthly, the ecological bias is a known and intrinsic limitation of meta regression analysis[33], since average patient characteristics are regressed against average trial outcomes and the data were extracted not at patients level[34]

## Conclusions

For each unitary increased of mTICI≥2b rate, the percentage of mRS<2 augment by 0.49 (CI95% 0.19-0.80) and the OR of obtaining a good functional outcome was 1.65 (CI95% 1.10-2.26), without significant differences between RCTs and registries performance.

## Data Availability

Since it is a systematic review, the data were deribed from literature

## Contributorship Statement

Conceptualization, GDR and EP; methodology, GDR, LP and CG; software, GDR and LP.; validation CG, EP, SF, SA and EC.; formal analysis, GDR and LP; investigation GDR, LB, SF, EP; resources LB; data curation, GDR, LB and LP.; writing—original draft preparation, GDR, EP; writing—review and editing CG; SA and EC.; visualization GDR, LP.; supervision, EP, CG, SF, SA EC;

## Competing Interests Statement

NA

All authors have read and agreed to the published version of the manuscript.

## Ethical approval

Not applicable, since this is a meta-regressions based on literature research

## Data sharing

Data are available upon request to the corresponding author

## References

1. Goyal M, Menon BK, van Zwam WH, et al. Endovascular thrombectomy after large-vessel ischaemic stroke: a meta-analysis of individual patient data from five randomised trials. Lancet 2016;387(10029):1723–31 doi: 10.1016/s0140-6736(16)00163-x[published Online First: Epub Date]|.

2. Broderick JP, Adeoye O, Elm J. Evolution of the Modified Rankin Scale and Its Use in Future Stroke Trials. Stroke 2017;48(7):2007–12 doi: 10.1161/STROKEAHA.117.017866[published Online First: Epub Date]|.

3. Powers WJ, Rabinstein AA, Ackerson T, et al. Guidelines for the Early Management of Patients With Acute Ischemic Stroke: 2019 Update to the 2018 Guidelines for the Early Management of Acute Ischemic Stroke: A Guideline for Healthcare Professionals From the American Heart Association/American Stroke Association. Stroke 2019;50(12):e344–e418 doi: 10.1161/STR.0000000000000211[published Online First: Epub Date]|.

4. Kaesmacher J, Dobrocky T, Heldner MR, et al. Systematic review and meta-analysis on outcome differences among patients with TICI2b versus TICI3 reperfusions: success revisited. Journal of Neurology, Neurosurgery & amp;amp; Psychiatry 2018;89(9):910 doi: 10.1136/jnnp-2017-317602[published Online First: Epub Date]|.

5. Dargazanli C, Fahed R, Blanc R, et al. Modified Thrombolysis in Cerebral Infarction 2C/Thrombolysis in Cerebral Infarction 3 Reperfusion Should Be the Aim of Mechanical Thrombectomy: Insights From the ASTER Trial (Contact Aspiration Versus Stent Retriever for Successful Revascularization). Stroke 2018;49(5):1189–96 doi: 10.1161/strokeaha.118.020700[published Online First: Epub Date]|.

6. Pego-Perez ER, Fernandez-Rodriguez I, Pumar-Cebreiro JM. National Institutes of Health Stroke Scale, modified Rankin Scale, and modified Thrombolysis in Cerebral Infarction as autonomy predictive tools for stroke patients. Reviews in the neurosciences 2019 doi: 10.1515/revneuro-2019-0011[published Online First: Epub Date]|.

7. Anglemyer A, Horvath HT, Bero L. Healthcare outcomes assessed with observational study designs compared with those assessed in randomized trials. Cochrane Database of Systematic Reviews 2014(4) doi: 10.1002/14651858.MR000034.pub2[published Online First: Epub Date]|.

8. Ross JS. Randomized Clinical Trials and Observational Studies Are More Often Alike Than Unlike. JAMA Internal Medicine 2014;174(10):1557–57 doi: 10.1001/jamainternmed.2014.3366[published Online First: Epub Date]|.

9. Deb-Chatterji M, Pinnschmidt H, Flottmann F, et al. Stroke patients treated by thrombectomy in real life differ from cohorts of the clinical trials: a prospective observational study. BMC neurology 2020;20(1):81 doi: 10.1186/s12883-020-01653-z[published Online First: Epub Date]|.

10. Ducroux C, Piotin M, Gory B, et al. First pass effect with contact aspiration and stent retrievers in the Aspiration versus Stent Retriever (ASTER) trial. 2020;12(4):386–91 doi: 10.1136/neurintsurg-2019-015215[published Online First: Epub Date]|.

11. Turk AS, 3rd, Siddiqui A, Fifi JT, et al. Aspiration thrombectomy versus stent retriever thrombectomy as first-line approach for large vessel occlusion (COMPASS): a multicentre, randomised, open label, blinded outcome, non-inferiority trial. Lancet 2019;393(10175):998–1008 doi: 10.1016/s0140-6736(19)30297-1[published Online First: Epub Date]|.

12. Yang P, Zhang Y, Zhang L, et al. Endovascular Thrombectomy with or without Intravenous Alteplase in Acute Stroke. New England Journal of Medicine 2020;382(21):1981–93 doi: 10.1056/NEJMoa2001123[published Online First: Epub Date]|.

13. Slim K, Nini E, Forestier D, et al. Methodological index for non-randomized studies (minors): development and validation of a new instrument. ANZ journal of surgery 2003;73(9):712–6 doi: 10.1046/j.1445-2197.2003.02748.x[published Online First: Epub Date]|.

14. Sterne JAC, Savović J, Page MJ, et al. RoB 2: a revised tool for assessing risk of bias in randomised trials. BMJ (Clinical research ed) 2019;366:4898 doi: 10.1136/bmj.l4898[published Online First: Epub Date]|.

15. Jackson D, White IR. When should meta-analysis avoid making hidden normality assumptions? Biometrical journal Biometrische Zeitschrift 2018;60(6):1040–58 doi: 10.1002/bimj.201800071[published Online First: Epub Date]|.

16. Clogg CC, Petkova E, Haritou A. Statistical methods for comparing regression coefficients between models. American journal of sociology 1995;100(5):1261–93

17. Manning NW, Chapot R, Meyers PM. Endovascular Stroke Management: Key Elements of Success. Cerebrovascular diseases 2016;42(3-4):170–7 doi: 10.1159/000445449[published Online First: Epub Date]|.

18. Andersson T, Wiesmann M, Nikoubashman O, et al. The Aspirations of Direct Aspiration for Thrombectomy in Ischemic Stroke: A Critical Analysis. J Stroke 2019;21(1):2–9 doi: 10.5853/jos.2018.02026[published Online First: Epub Date]|.

19. Frieden TR. Evidence for Health Decision Making - Beyond Randomized, Controlled Trials. The New England journal of medicine 2017;377(5):465–75 doi: 10.1056/NEJMra1614394[published Online First: Epub Date]|.

20. Spieth PM, Kubasch AS, Penzlin AI, et al. Randomized controlled trials - a matter of design. Neuropsychiatric disease and treatment 2016;12:1341–9 doi: 10.2147/NDT.S101938[published Online First: Epub Date]|.

21. Faraoni D, Schaefer ST. Randomized controlled trials vs. observational studies: why not just live together? 2016;16(1):102

22. Mansournia MA, Higgins JP, Sterne JA, et al. Biases in Randomized Trials: A Conversation Between Trialists and Epidemiologists. Epidemiology 2017;28(1):54–59 doi: 10.1097/EDE.0000000000000564[published Online First: Epub Date]|.

23. Schnell-Inderst P, Iglesias CP, Arvandi M, et al. A bias-adjusted evidence synthesis of RCT and observational data: the case of total hip replacement. Health Economics 2017;26(S1):46–69 doi: https://doi.org/10.1002/hec.3474[published Online First: Epub Date]|.

24. Gamerman V, Cai T, Elsäßer A. Pragmatic randomized clinical trials: best practices and statistical guidance. Health Services and Outcomes Research Methodology 2019;19(1):23–35 doi: 10.1007/s10742-018-0192-5[published Online First: Epub Date]|.

25. Saturni S, Bellini F, Braido F, et al. Randomized controlled trials and real life studies. Approaches and methodologies: a clinical point of view. Pulmonary Pharmacology & Therapeutics 2014;27(2):129–38 doi: https://doi.org/10.1016/j.pupt.2014.01.005[published Online First: Epub Date]|.

26. Chalos V, van der Ende NAM, Lingsma HF, et al. National Institutes of Health Stroke Scale: An Alternative Primary Outcome Measure for Trials of Acute Treatment for Ischemic Stroke. Stroke 2020;51(1):282–90 doi: 10.1161/STROKEAHA.119.026791[published Online First: Epub Date]|.

27. Huang H-C, Tsai J-Y, Liu T-C, et al. Functional recovery of stroke patients with postacute care: a retrospective study in a northern medical center. Journal of the Chinese Medical Association 2019;82(5)

28. Deutsch A. Does Postacute Care Site Matter? A Longitudinal Study Assessing Functional Recovery After a Stroke. Archives of Physical Medicine and Rehabilitation 2013;94(4):630–32 doi: https://doi.org/10.1016/j.apmr.2013.02.011[published Online First: Epub Date]|.

29. Middleton S, McElduff P, Ward J, et al. Implementation of evidence-based treatment protocols to manage fever, hyperglycaemia, and swallowing dysfunction in acute stroke (QASC): a cluster randomised controlled trial. The Lancet 2011;378(9804):1699–706

30. Cramer SC, Wolf SL, Adams Jr HP, et al. Stroke recovery and rehabilitation research: issues, opportunities, and the National Institutes of Health StrokeNet. Stroke 2017;48(3):813–19

31. Berkhemer OA, Fransen PS, Beumer D, et al. A randomized trial of intraarterial treatment for acute ischemic stroke. The New England journal of medicine 2015;372(1):11–20 doi: 10.1056/NEJMoa1411587[published Online First: Epub Date]|.

32. Nogueira RG, Haussen DC, Castonguay A, et al. Site Experience and Outcomes in the Trevo Acute Ischemic Stroke (TRACK) Multicenter Registry. Stroke 2019;50(9):2455–60 doi: 10.1161/STROKEAHA.118.024639[published Online First: Epub Date]|.

33. Baker WL, White CM, Cappelleri JC, et al. Understanding heterogeneity in meta-analysis: the role of meta-regression. International journal of clinical practice 2009;63(10):1426–34 doi: 10.1111/j.1742-1241.2009.02168.x[published Online First: Epub Date]|.

34. Berlin JA, Santanna J, Schmid CH, et al. Individual patient-versus group-level data meta-regressions for the investigation of treatment effect modifiers: ecological bias rears its ugly head. Statistics in medicine 2002;21(3):371–87

